# A Large Language Model-Based Generative Natural Language Processing Framework Finetuned on Clinical Notes Accurately Extracts Headache Frequency from Electronic Health Records

**DOI:** 10.1101/2023.10.02.23296403

**Authors:** Chia-Chun Chiang, Man Luo, Gina Dumkrieger, Shubham Trivedi, Yi-Chieh Chen, Chieh-Ju Chao, Todd J. Schwedt, Abeed Sarker, Imon Banerjee

## Abstract

**Background:** Headache frequency, defined as the number of days with any headache in a month (or four weeks), remains a key parameter in the evaluation of treatment response to migraine preventive medications. However, due to the variations and inconsistencies in documentation by clinicians, significant challenges exist to accurately extract headache frequency from the electronic health record (EHR) by traditional natural language processing (NLP) algorithms.

**Methods:** This was a retrospective cross-sectional study with human subjects identified from three tertiary headache referral centers-Mayo Clinic Arizona, Florida, and Rochester. All neurology consultation notes written by more than 10 headache specialists between 2012 to 2022 were extracted and 1915 notes were used for model fine-tuning (90%) and testing (10%). We employed four different NLP frameworks: (1) *ClinicalBERT (Bidirectional Encoder Representations from Transformers) regression model* (2) Generative Pre-Trained Transformer-2 (*GPT-2) Question Answering (QA) Model zero-shot (3) GPT-2 QA model few-shot training* fine-tuned on Mayo Clinic notes; and *(4) GPT-2 generative model few-shot training* fine-tuned on Mayo Clinic notes to generate the answer by considering the context of included text.

**Results:** The GPT-2 generative model was the best-performing model with an accuracy of 0.92[0.91 – 0.93] and R^2^ score of 0.89[0.87, 0.9], and all GPT2-based models outperformed the ClinicalBERT model in terms of the exact matching accuracy. Although the ClinicalBERT regression model had the lowest accuracy 0.27[0.26 – 0.28], it demonstrated a high R^2^ score 0.88[0.85, 0.89], suggesting the ClinicalBERT model can reasonably predict the headache frequency within a range of ≤ ± 3 days, and the R^2^ score was higher than the GPT-2 QA zero-shot model or GPT-2 QA model few-shot training fine-tuned model.

**Conclusion:** We developed a robust model based on a state-of-the-art large language model (LLM)-a GPT-2 generative model that can extract headache frequency from EHR free-text clinical notes with high accuracy and R^2^ score. It overcame several challenges related to different ways clinicians document headache frequency that were not easily achieved by traditional NLP models. We also showed that GPT2-based frameworks outperformed ClinicalBERT in terms of accuracy in extracting headache frequency from clinical notes. To facilitate research in the field, we released the GPT-2 generative model and inference code with open-source license of community use in GitHub.

## Introduction

Headache medicine is a unique field where the diagnosis, treatment, and outcome assessments rely heavily on human natural language rather than test results. Important clinical information for diagnosis and evaluation of treatment responses, including detailed headache description, migraine-associated symptoms, aura, prior treatment trials, and the frequency and severity of migraine attacks and headaches are often documented as free text in clinical notes in the Electronic Health Records (EHRs). Migraine is a highly prevalent and disabling neurological condition that affects around 16% of the United States population and more than 1 billion people worldwide, causing significant disability and loss of productivity[1]. For patients with frequent migraine attacks, preventive therapeutic interventions are recommended with the aim of reducing their headache frequency and severity. Currently, the gold standard metric for evaluation of treatment response to migraine preventive medications is the change in headache frequency, often documented as headache days per month or every four weeks in clinicians’ notes. Varying syntactic reporting of headache frequency poses significant challenges when large-scale extraction of such data is needed, and conducting chart reviews by humans reading the notes is time-consuming and not practical when a large amount of data is needed for research.

Natural Language Processing (NLP) is a branch of Artificial Intelligence (AI) that concerns the ability of machines to understand, interpret, and generate human language. Previous attempts have used NLP to extract headache-related information from various sources to perform tasks including analyzing patient self-written narratives to distinguish between migraine and cluster headache[2], developing a generalizable NLP model to identify users with self-reported migraine on various social media platforms[3], and distinguishing between migraine versus other headache as well as identifying headache-associated symptoms[4]. Although headache frequency is one of the most commonly used gold standard metrics in the evaluation of treatment responses, currently, there are no tools reported to accurately and reliably extract headache frequency from various clinic notes within the EHR. Possible explanations include 1) Headache frequency is not always documented for all patients being evaluated for migraine, especially when patients were seen by non-specialists; 2) Even if the information exists, there is a lack of consistency in the documentation of headache frequency, making it challenging to extract such information based on simple rule-based NLP models. Here we list several examples of variations of documentation of headache frequency in clinical notes, defined as days with any headache in a 4-week period (28 days).*“She experiences daily headache with 10 severe migraine days per month”*: it requires the recognition of “daily” headache (e.g., 28 days per month with headache) instead of picking up the number 10, *“He reports having headaches 3 days per week”*-requires calculation 3 x 4 weeks= 12 days, and *“Overall, she has 2 headache-free days per month”*-requires calculation 28-2=26 days.

With the advances in NLP techniques, the recent emergence of generative large language models (LLMs), and the confluence of AI, there arises an opportunity to elevate the granularity and accuracy of automated data extraction from EHRs. We aim to develop robust modeling frameworks that can accurately extract headache frequency data from free-text clinical notes in the EHR. We performed a comparative analysis of a traditional transformer-based NLP model, notably a ClinicalBERT regression model, against the recent advancement of open source, generative LLM models, namely three Generative Pre-Trained Transformer-2 (GPT-2) based models with various architectures - (a) zero-shot versus few-shot; (b) question-answering versus generative. We hypothesized that novel LLM frameworks could overcome the challenges in headache frequency extraction that would be otherwise hard to achieve with traditional transformer-based NLP models.

## Methods

This was a retrospective cross-sectional study with human subjects identified from Mayo Clinic in Arizona, Florida, and Rochester. The Mayo Clinic Institutional Review Board approved an exemption for this study, and written informed consent was not required. We extracted 34369 neurology consultation notes documented by more than 10 headache specialists within 2012 – 2022 across 3 geographically distributed sites and tertiary referral Headache Centers in Rochester, MN, Scottsdale, AZ, and Jacksonville, FL. We randomly selected 1,950 notes for model development. After excluding irrelevant notes, we curated 1,915 notes for model fine-tuning (90%) and testing (10%). Three individual readers annotated the headache frequency from the curated cohort. All headache frequency annotations were validated by headache specialists (CC and TS) to ensure accuracy and used as the gold standard for model development.

### Experimental Design

Figure 1 shows the overall framework for our experimental design. After Name Entity Recognition (NER) extraction, four different state-of-the-art NLP modeling architectures were used to directly read the full neurology consultation notes selected for model development and to extract documented headache frequencies. We then compare the performance metrics against the manual annotations confirmed by headache specialists. The subsections below detail each component of the modeling framework including a brief description of the model architecture variations.

**Fig 1.**
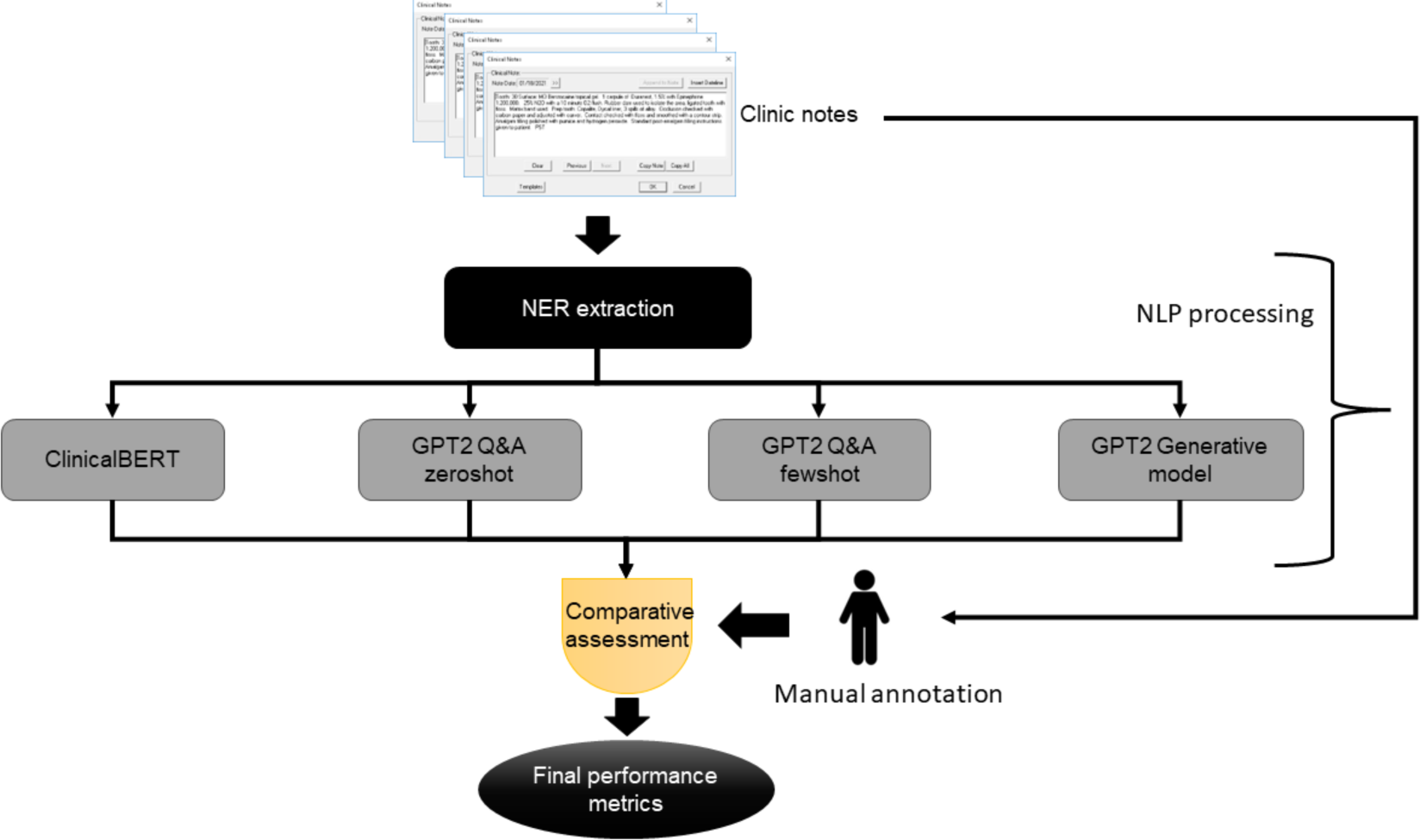
Nature Language Processing (NLP) framework for extracting headache frequency from neurology consultation notes - four parallel modeling schemes: (1) *ClinicalBERT regression model*: encoder-based regression model pre-trained on MIMIC-III; (2) *GPT-2 QA model zero-shot:* decoder QA model trained on the generic web scraped data; *(3) GPT-2 QA model few-shot training:* decoder model trained on the generic web scraped data and fine-tuned on Mayo Clinic notes; *and (4) GPT-2 generative model few-shot training:* decoder model trained on the generic web scraped data and fine-tuned on Mayo Clinic notes to generate directly the answer by considering the context text. Abbreviations: Name Entity Recognition (NER); The Medical Information Mart for Intensive Care III (MIMIC-III); Generative Pre-Trained Transformer-2 (GPT-2); Question Answering (QA)

### NER extractions

To capture the vocabulary for the intended task, we compiled the following two complementary dictionaries: the target term list, which was a publicly available terminology program (Clinical Event Recognizer) extended with 10 additional terms for migraine that were primarily captured by analyzing the training set, including migraine, headache, headaches, days per month, frequency; and the modifier list, which was a list of modifier terms, including clinical terms related to negations (eg, no, rule out), temporality (eg, history, current), family (eg, mother, sister) and discussion (eg, risk of, may introduce). Finally, a keyword-based sentence retrieval method was applied to each clinic note, which selected only the sentences that contained at least one of the migraine/headache-related terms as a named entity and generated a text snippet by combining the sentences extracted from the whole notes. Based on the modifiers, we dropped all the historical reporting and discussion.

### Language modeling

In parallel, we compared four state-of-the-art strategies for extracting headache frequency from the selected text block - (1) *ClinicalBERT regression model*: an encoder-based regression model pre-trained on the Medical Information Mart for Intensive Care III (MIMIC-III), which is a large database with deidentified health-related data associated with over 40,000 patients who stayed in critical care units of the Beth Israel Deaconess Medical Center between 2001 and 2012 [5]; (2) *GPT2-Question Answering (QA) Model zero-shot:* a decoder model trained on the generic web scraped data to <u>predict relevant answer text position within the context text</u>; (3) *GPT-2 QA model few-shot training:* a decoder model trained on the generic web scraped data and fine-tuned on the selected 1,915 Mayo Clinic neurology consultation notes with annotations confirmed by headache specialists to <u>predict relevant answer text position within the context text</u>; and (4) *GPT-2 generative model few-shot training:* a decoder model trained on the generic web scraped data and fine-tuned on the same 1,915 Mayo Clinic notes to <u>generate the answer directly by considering the context of the included text which does not require the answer to be included in the sentences used</u>. We chose GPT-2 based models as those are open-source models that can be downloaded locally and fine-tuned as needed without the risk of uploading sensitive information to a third party.

### ClinicalBERT regression model

ClinicalBERT[6] has a bidirectional encoder representation similar to the BERT, where ClinicalBERT is pre-trained on clinical data - MIMIC-III clinic notes, and BERT is trained on generic domain - Wikipedia and BookCorpus. Both models are designed to understand the context of words in a sentence by looking at the words that come both before and after each word in a sentence. They are trained via masked language modeling (MLM) - an unsupervised learning approach that can utilize large amounts of unannotated free text. MLM training masks some random words in the input and the model is trained to predict missing words in a sentence. We fine-tuned ClinicalBERT and trained a regression model. More specifically, given an input sentence, we used ClinicalBERT to encode the sentence into a sequence of tokens starting with a special token [CLS][7], Similar to any other BERT model, such that the [CLS] model is a global representation of the input sentence. During the fine-tuning phase of the model, we initialized the model with the weights of a pre-trained model “emilyalsentzer/Bio_ClinicalBERT”[6,8], which is pre-trained on MIMIC-III corpus. On top of ClinicalBERT, we formulated the headache frequency detection as an ordinal regression problem where each input text will obtain a score with the interval [-1,28], -1 being no headache frequency detected in the note and 28 indicating having daily headache, using 28 days (4 weeks) as a unit to capture headache days per month. We use mean square loss as the objective function to train the regressor. The learning rate is set 5e-5, and total training epoch is 50, batch size of 5. The optimizer chosen for this task was AdamW[9,10]

### GPT2-QA Model zero-shot and few-shot training

GPT-2 is a decoder-only architecture, which is an autoregression model. The main difference between the autoregression and bidirectional encoder, like the BERT model, is that the autoregressive model generates sentences word by word, each time predicting the next word based on all the previous words in the sentence rather than both previous and following words as in the bidirectional model. Leveraging unsupervised training strategy, GPT-2 is trained on a large training corpus of web-scraped data but with the next word prediction task, which means the model is trained to predict the next word in a sentence given all the previous words. The GPT-2 model is good at text generation tasks given the nature of how it was pre-trained. We frame GPT-2 as an extractive QA model. For such a QA task, the model is given a context and a question, and the goal is to extract the answer to the question from the context.

To extract the answer, the GPT-2 QA first tokenizes an input sentence into a sequence of tokens. Every token is represented by a vector embedding. Then the model predicts which token is the start token of an answer and which token is an end token of the answer. To achieve this, on top of the GPT-2 model, we add a linear layer that takes each token as input and predicts two probabilities of this token: being the start token of the answer and being the end token of the answer. Finally, the token that has the highest probability of being the start token is the beginning of the answer, like the end token of the answer. Considering the following example, the context is “Over the past four weeks, she reports having had 15 headache days.”, and the question is “*what is the monthly frequency of the headache for this patient?*” The label of this example is (11, 12) since token 11 and token 12 represent the answer “15” in the context. During the training, the model is trained to predict the highest probability (i.e. label 1) of token 11 to be the start token, and the low probabilities of every other token (label 0). Similarly, token 12 has the highest probability of being the end (label 1) and the probability of being the end token for every other token is low (label 0). We use cross entropy loss as the training objective.

We initialize the model with the pretrained weights of “anas-awadalla/gpt2-span-head-few-shot-k-32-finetuned-squad-seed-4”, which is fine-tuned on the Squad dataset[11]. The learning rate is set 2e-5, and total training epoch is 10, batch size of 16. The optimizer chosen for this task was AdamW[9,10].

First, we perform a zero-shot testing of the QA model “anas-awadalla/gpt2-span-head-few-shot-k-32-finetuned-squad-seed-4”. Since the model is trained on a question-answering task before, theoretically, we can use it to answer any question. One concern is the domain shift issue which characterizes the change in statistical distribution or semantic organization of the data, e.g. the model trained on politics, is prompted to answer questions regarding clinical problems; however, our target task does not require any clinical or biomedical background to answer the question, thus, we hypothesize that the QA model can do reasonable zero-shot prediction. In this zero-shot evaluation setting, we provided the question: “*What is the headache frequency per month?*’” to test the pre-trained QA model. From our analysis, which will be detailed below, we found that while the exact matching score is not high, the model was able to predict a span which contained the answer. For example, the ground truth is “12”, and the model generates an answer “a headache for 12 days out of 28”; another example: the ground truth is “28”, and the model generates an answer “a daily headache”. Semantically, the model’s prediction is correct; however, by exact matching, the model will be judged as incorrect. To resolve this evaluation problem and fair comparison, we mapped the zero-shot model prediction to a digit answer by extracting the digit value in the prediction, or if the words “daily” or “every day” mentioned in the prediction, we map the prediction to “28”; if there is no digit mentioned in the prediction, we map the prediction to “-1”.

In addition, we have applied a few-shot learning strategy where we showed some examples to the GPT-2 QA model for fine-tuning the answer space to extract more targeted answers as expected by the experts. We fine-tuned the same model using the 1,915 annotated notes and reported the performance on the test set.

### GPT-2 Generative Model

The GPT-2 QA model assumes that the answer is always a span in the input text; however, it fails when the answer is not directly given in the input. For example, the given text can be “*the patient has 4 headache-free days a month.*”, then the actual answer is 28-4=24 days which is not present within the context. To resolve such a challenge, we train a GPT-2 generative model to generate the actual frequency instead of the index of the answer in the given text. Furthermore, since the possible answers are discrete values ranging from -1 to 28, where -1 represents there is no frequency mentioned in the given text, we add all the possible answers, all numbers from -1 to 28, as new tokens to the model’s vocabulary if they are not already part of it. During the training, the model is optimized by negative log-likelihood of the frequency label. Here, we use the same example given in the previous section to illustrate the main difference between the extractive QA counterpart model. Given the same context and the question, the label for the GPT-2 generative model is “15”. On top of GPT-2, we add a linear layer with the embedding as input and cross entropy loss as the training objective. To fairly compare with the GPT-2 QA model, we initialize the GPT generative model with the same pretrained weights and same hyperparameters during the training time.

## Results

### Cohort

A total of 1,915 neurology consultation notes written by headache specialists were used for model development. Three individual readers extracted the headache frequency from the curated cohort and inter-rater reliability was >0.85.

### Quantitative performance

We compared the model extracted frequencies against the manual labeled ground truth and reported the performance for all four models in **Table 1**. Overall, the GPT-2 generative model was the best-performing model, with an accuracy of 0.92 [0.91 – 0.93] and R^2^ score of 0.89 [0.87, 0.9], followed by the GPT-2 QA model (accuracy 0.87 [0.85, 0.87], R^2^ 0.53 [0.45, 0.55]), GPT-2 zero-shot model (accuracy 0.57 [0.57, 0.59], R^2^ -0.014 [-0.81, -0.01]), then the ClinicalBERT regression model (accuracy 0.27 [0.26 – 0.28], R^2^ 0.88 [0.85, 0.89]), though the ClinicalBERT model demonstrated a high R^2^ score.

**Table 1.** Quantitative performance of the parallel architectures. Optimal performance is highlighted in bold. 95% confidence interval is calculated using auto-bootstrapping and represented as [x, y]. Same test set is used for all the model evaluations.

| Model | Accuracy | R2 score |
| --- | --- | --- |
| ClinicalBert regression | 0.27 [0.26, 0.28] | **0.88 [0.85, 0.89]** |
| GPT2 zeroshot | 0.57 [0.57, 0.59] | -0.014 [ -0.81, -0.01] |
| GPT2 QA model | 0.87 [0.85 , 0.87] | 0.53 [0.45, 0.55] |
| GPT2 generative model | **0.92 [0.91, 0.93]** | **0.89 [0.87, 0.9]** |

The results show that GPT-2 type models outperformed the ClinicalBERT model in terms of the exact matching accuracy. Notably, even the zero-shot QA model performed significantly better than the ClinicalBERT model in terms of classification accuracy. This suggests that classifying one label from 30 classes (i.e. discrete value of [-1, 28]) in ClinicalBERT is not an effective approach for our task. In contrast, the R^2^ score of ClinicalBERT is high, which suggests that the ClinicalBERT model can predict the label within a reasonable range (≤ ± 3) but the GPT-2 QA fine-tuned model only scored 0.53 in R^2^ score. Comparing the GPT-2 QA and GPT-2 generative models, the latter performs better than the QA model in terms of both scores. The advantage of generative models is that when the answer directly appears in the text, the generative model can still do well, but the extractive QA model might fail.

In order to further analyze the model performance, we visualized the scatter plot of true and predicted values in Fig. 2 and Bland-Altman plot in Fig. 3. GPT-2 zero-shot, even after post-processing, is under-estimating the migraine frequency (mean -7.15) and not able to extract the frequency if it is not reported directly using simple language. As seen in Figures 2 and 3, the fine-tuned GPT-2 QA model trained with cross-entropy loss is able to estimate the frequency with moderate alignment and mean difference is 2.37 (+/-12 1.96 std), while fine-tuned ClinicalBERT achieved decent alignment and mean difference is 0.78 (+/-6.6 1.96 std). However, the GPT-2 generative model outperformed all the architecture and achieved 0.46 mean difference (+/- 7.1 1.96 std).

**Fig 2.**
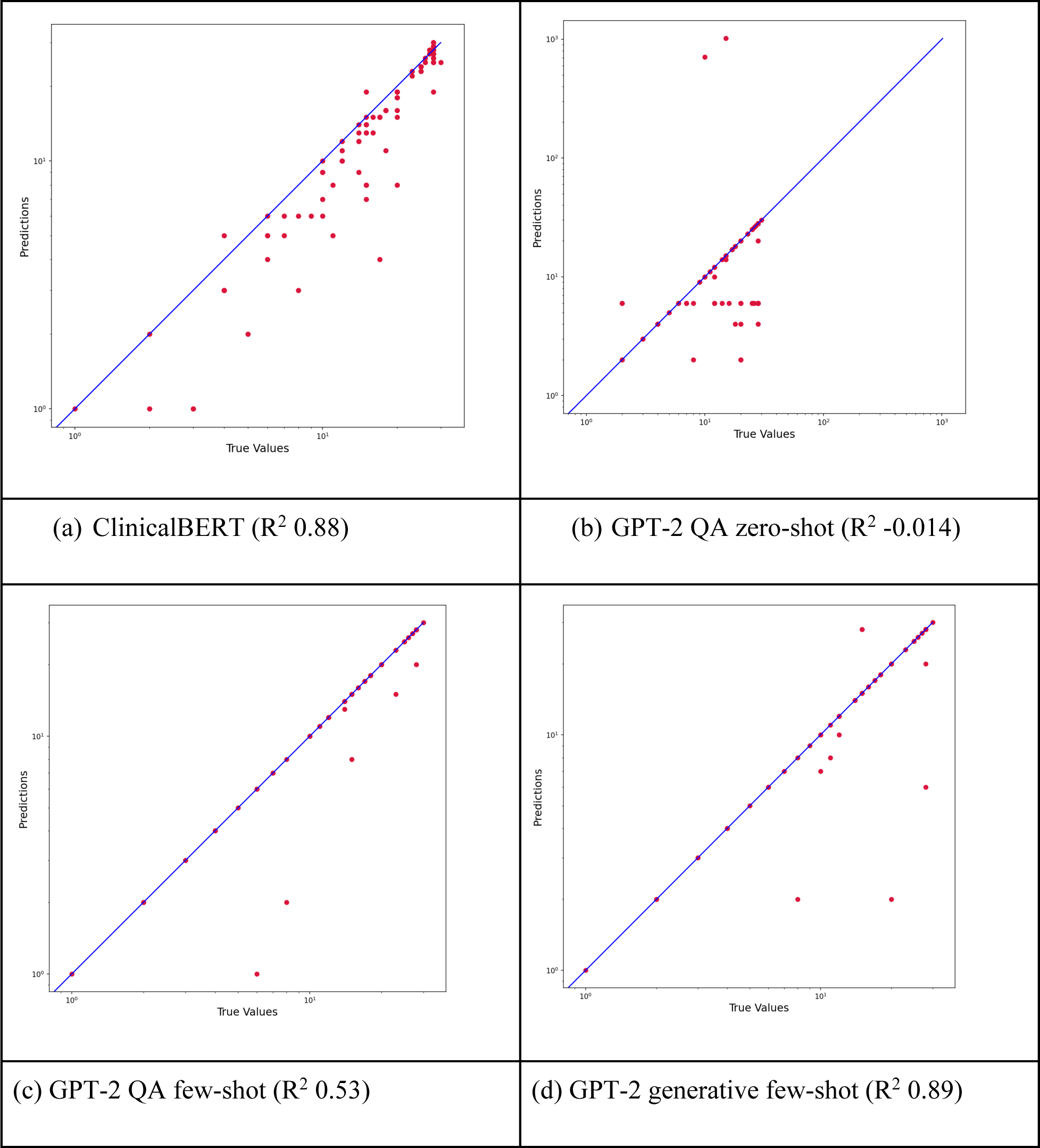
Scatter plot visualization of true frequency reported in clinic notes (x-axis) and predicted frequency by the model (y-axis) in log scale. Blue line shows the perfect alignment (R^2^ = 1.0).

**Fig 3.**
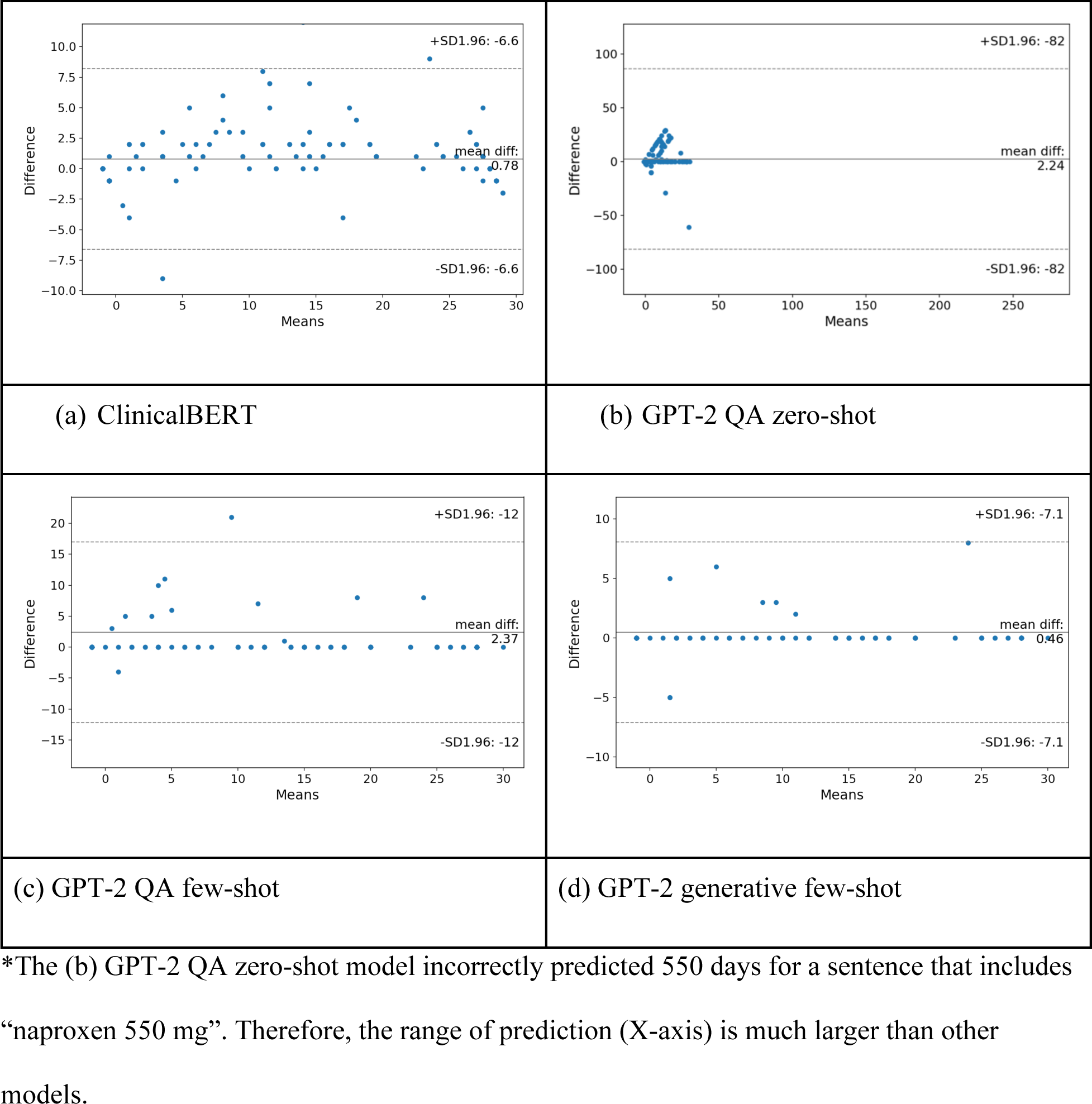
Bland-Altman test to compare each metric computed from the NLP model against the ground truth. There are 192 data elements in total for each subfigure, with each point representing one note in the validation dataset. Mean and standard deviation is also calculated.

### Error Analysis

While the overall performance is 92% accuracy, the GPT-2 generative model still makes mistakes on 8% of cases when more complicated reasoning is required. For example, *“The patient generally has two headache-free days per week”* The correct answer would be (7-2) x 4 = 20; however, the model predicted 2 days per month. When headache frequency is documented as a range, we labeled the maximum frequency as the ground truth, while the model often extracts the minimum headache frequency. For example, ’On average, she estimates 7-10 headache days per month.’ The model reports 7 days which was also correct, while the ground truth label was 10 days. More examples are presented in Table 2.

**Table 2.**
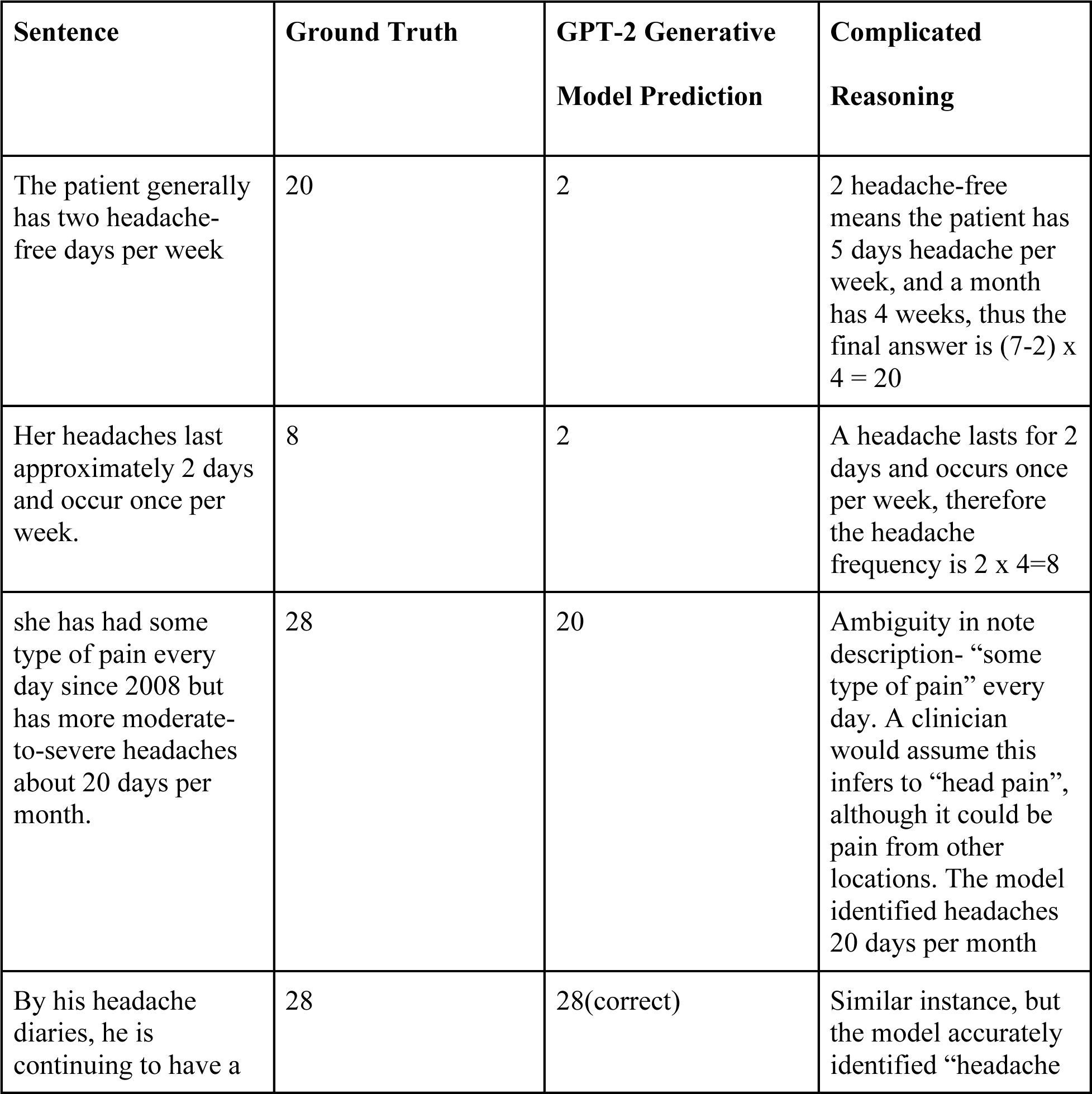
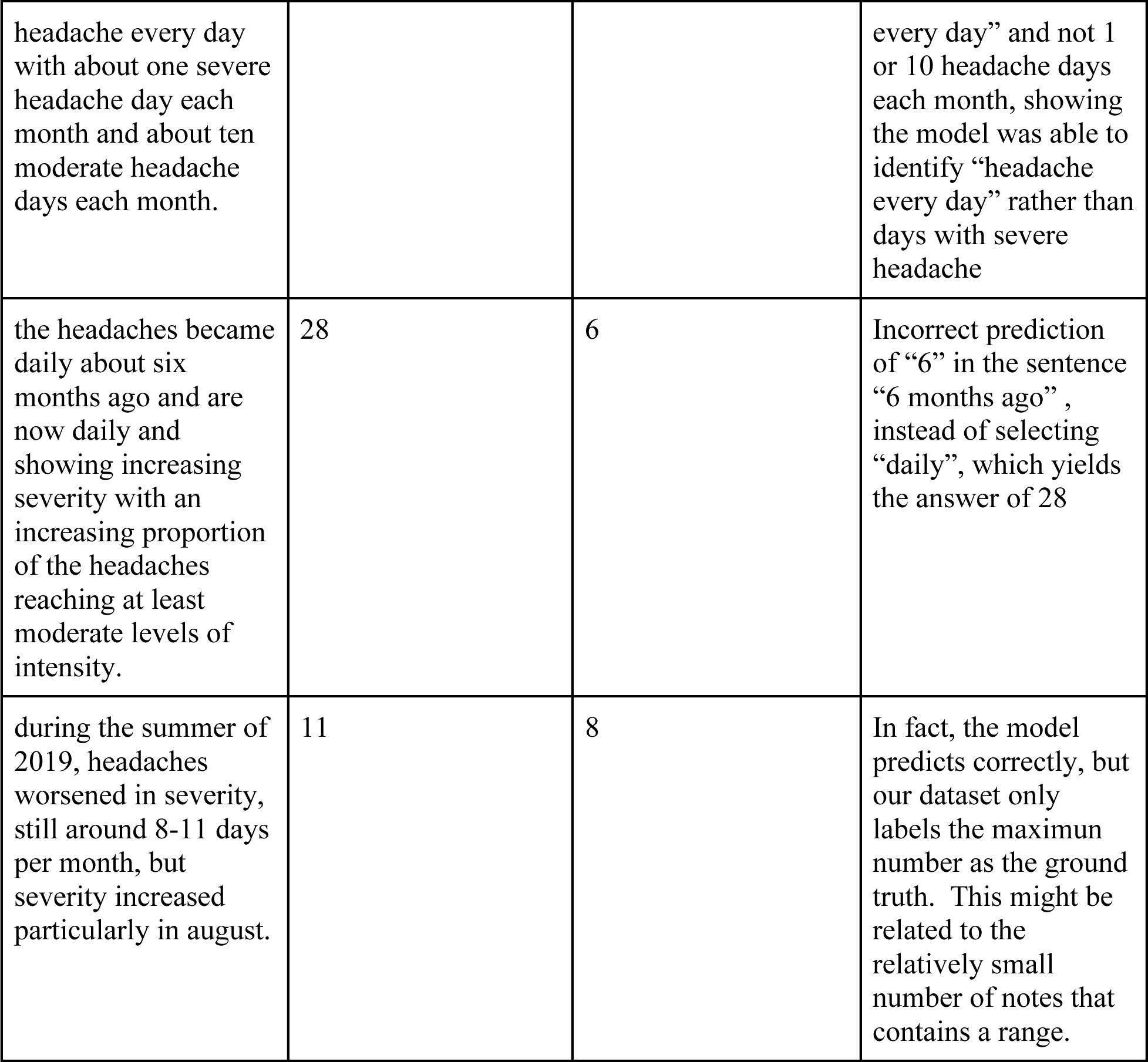
Example cases selected for error analysis; We provide examples of errors from the GPT-2 generated model and also highlight the reasoning for analysis.

## Discussion

We experimented with several state-of-the-art models that can reliably extract headache frequency (monthly headache days) based on clinical notes in the EHR with high performance and showed that GPT-2 based models, specifically the GPT-2 generative model, with an accuracy of 0.92 [0.91 – 0.93] and R^2^ score of 0.89 [0.87, 0.9], outperformed the traditional ClinicalBERT regression model (accuracy 0.27 [0.26 – 0.28], R^2^ 0.88 [0.85, 0.89]). Even though ClinicalBERT had lower accuracy, it achieved a high R^2^ score, indicating it can reasonably predict headache frequency for a given text within a reasonable range (≤ ± 3).

While the GPT-2 zero-shot model did not perform well (accuracy 0.57 [0.57, 0.59], R^2^ -0.014 [- 0.81, -0.01]), the results are decent given that no training data is needed for the model. This result is aligned with the literature where researchers found that GPT-2 is a good zero-shot learner[12]. We applied few-shot in-context learning with the same GPT-2 model which improves the performance from the zero-shot (57% - 87% accuracy) (accuracy 0.87 [0.85, 0.87], R^2^ 0.53 [0.45, 0.55] for the GPT-2 QA model), but it failed to extract the headache frequency when the exact answer is not present and/or additional reasoning is required.

In developing the model to extract headache frequency documented in clinical notes, we were able to overcome challenges related to inconsistencies in documenting headache frequencies, which can present in various forms in the note. While sentences like *“Over the past 4 weeks, he experiences 13 headache days per month”,* and *“Headache frequency: 26 days’’* are straightforward for all models to extract the number documented in the note, we noticed the ClinicalBERT model could not reliably capture and often underestimates descriptions of daily or constant headache, which represents the majority of patients seen at tertiary headache centers. Examples include *“She continues to experience constant headaches*”-answer is 28, and *“His headache is still constant and daily with severe headaches occurring on 10 days per month*”-answer is 28 and not 10. We overcame these challenges by fine-tuning the LLM models QA and generative paradigm by expert-annotated clinic notes, and the model learned the patterns and generated the correct answers.

In other instances, the accurate answer does not exist within the sentence and requires calculation based on the content of the sentence. For example, *“She has 5 headache-free days per month-answer is 28-5=23”,* and *“On average, she has 4 headache days per week-answer is 4x4=16”*. We were able to overcome those challenges by employing a GPT-2 generative model to comprehend, calculate and generate the answer instead of extracting the index of the answer in the given text.

Although previous studies have utilized NLP to extract migraine characteristics from the EHR[2,4], accurate extraction of headache frequency has been a challenging task, yet of great importance. To our knowledge, this is the first study that reports NLP frameworks that can accurately extract headache frequency using LLMs. Extraction of headache frequency is of particular importance since change in headache frequency is a commonly used measure for determining the effectiveness of migraine preventive treatment. The strengths of our study include employing and comparing several LLM-based models with different training architectures-QA versus generative, and strategies-zero-shot versus few-show inference to overcome the challenges of variation in clinical documentation of headache frequency. We achieved high accuracy and R^2^ score with the GPT-2 generative model. Even though we were able to achieve 92% accuracy in extracting headache frequency, the study has several limitations. First, the models were fine-tuned and tested only on clinic notes written by headache specialists at Mayo Clinic. While the test set includes notes documented by more than 10 different headache specialists from different sites (Rochester, Arizona, and Florida) with various syntactic documentation patterns and preferences, the model may need additional fine-tuning for an outside organization with different practice patterns and patient populations. Additionally, the model has only been tested on the neurology consultation notes. Extracting headache frequency documented by other specialties or from data sources other than clinic notes might require additional fine-tuning of the model.

## Conclusion

We leverage the generative capacity of LLM and developed high-performing, state-of-the-art models with high accuracy and R^2^ score to accurately extract headache frequency documented in clinical notes in the EHR. We showed that GPT-2 based models outperformed traditional transformer model-ClinicalBERT regression, and compared different LLM-based frameworks with GPT-2 zero-shot, few-shot, question answering model, and generative models. Our results showed that the GPT-2 generative model was the best-performing model that could recognize various ways of describing headache frequency in clinical notes. We developed a powerful tool for EHR-based headache research as changes in headache frequency remain the gold standard for evaluation of treatment outcomes to migraine preventive medications. To facilitate research in the field, we released the GPT-2 generative model and inference code with open-source license of community use in GitHub. https://github.com/imonban/MigraneFreq_extract/tree/main

### Funding source

This study is partially supported by NIH/NCI, U01 CA269264-01-1, Flexible NLP toolkit for automatic curation of outcomes for breast cancer patients. This research was funded in part by the Fisher Benefactor Fund for Migraine and Headache Research at Mayo Clinic.

### Financial Disclosures

In the prior 24 months, CC has received personal compensation for consulting with Satsuma and eNeura. She also receives research support from American Heart Association with grants paid to her institution.

In the prior 24 months, TJS has received personal compensation for consulting with Abbvie, Allergan, Amgen, Axsome, Biodelivery Science, Biohaven, Collegium, Eli Lilly, Linpharma, Lundbeck, Satsuma, and Theranica; research grant support from American Heart Association, American Migraine Foundation, Amgen, Henry Jackson Foundation, NIH, PCORI, SPARK Neuro, United States Department of Defense; royalties from UpToDate; stock options from Aural Analytics and Nocira.

Other authors report no financial disclosure.

## Data Availability

All data produced in the present study are available upon reasonable request to the authors

## Notes

### Competing Interest Statement

The authors have declared no competing interest.

### Author Declarations

The Mayo Clinic Institutional Review Board approved an exemption for this study, and written informed consent was not required.

